# A retrospective analysis of COVID-19 non-pharmaceutical interventions for Mexico and Peru: a modeling study

**DOI:** 10.1101/2022.12.19.22283668

**Authors:** M. Adrian Acuña-Zegarra, Mario Santana-Cibrian, Carlos E. Rodriguez Hernandez-Vela, Ramsés H. Mena, Jorge X. Velasco-Hernández

## Abstract

We model the observed dynamics of COVID-19 in Mexico and Peru and explore the impact of hypothetical non-pharmaceutical interventions applied on key days of civic, religious, or political nature that increased contacts and transmission events. Using as a baseline the observed epidemic curve, we apply hypothetical reductions in the contact rates during the first year of the pandemic: i) near the beginning, ii) at the beginning of the second outbreak, and iii) end of the year. The effects of the interventions are different for Mexico and Peru but underlie the fact that strong early interventions do reduce the prevalence and, in general, allow for an epidemic evolution of relatively lower prevalence than interventions applied once the epidemic is underway. We provide evidence that key calendar days are good approximations of times when contact rates change and, therefore, are efficient periods for effective interventions particularly in places with low testing and lack of contact tracing. This has helped us to recreate different outbreaks of the COVID-19 disease dynamics in Mexico and Peru and explore the impact of hypothetical interventions that reduce the contact rate.

## 1. Introduction

Emerging infectious diseases can be defined as infections that have newly appeared in a population or have existed but are rapidly increasing in incidence or geographic range [24]. Recent examples are H1N1 influenza (2009), Chikungunya (2014), Zika (2015), and COVID-19 (2019 to the present), the latter being the cause, so far, of more than 6.6 million deaths around the world [22].

Mathematical modeling has been widely used to study the COVID-19 epidemic. The transmission dynamics has been described with many different methodologies, several of them centered on estimating the effective reproduction number *R*_*t*_ with some version of the well-known Kermack-McKendrick model [7, 17, 30, 35]. During 2020 and 2021, much effort was centered on projecting the COVID-19 pandemic and evaluating the efficacy of the mitigation strategies adopted to contain it [6, 16]. Around the world, the implementation of these measures has varied in strength ranging from strict and mandatory governmental enforcement to a voluntary personal decision. Regardless of the particular version of the mitigation strategy followed, these strategies were based on local factors that combine public health status, economic impact and political conditions [4].

The central phenomena of human behavior that we explore are superspreading events occurring in particular calendar dates associated with religious, commercial, or civic holidays specific to each country. During or after these events (depending on their length), the contact rate changes. We, therefore, use these events as change points. We argue that in Mexico and Peru (both middle-income countries with a very stressed economic activity due to the pandemic), these change points (key calendar dates) reflect on the contact rate more clearly than changes imposed by the government (non-pharmaceutical interventions or NPIs) [13, 19]. Using key calendar dates has another advantage from the perspective of forecasting: these dates are known in advance and therefore mobility can be anticipated (short vacations, family visits, buying sprees, etc).

Here, we present a methodology that uses the known history of the disease reflected in the contact rate as a baseline to recreate the observed disease dynamics. In [13], the authors have used a similar idea but, in our case, to determine changes in the contact rate, we look at particular events occurring on dates related to school, civic or religious periods (vacations, civic holidays, commercial events) that are known in advance each year for a given country. With this historical information in hand, we describe the short-term evolution of the COVID-19 pandemic. Our results are illustrated considering as examples some Mexican states (Mexico City, Queretaro, Quintana Roo, and Sonora) and some departments of Peru (Arequipa, Cusco, Lima, and Piura). The main objective is to analyze, in retrospective, the impact of hypothetical non-pharmaceutical interventions in several keydates. In particular, we analyze the effect of these interventions in the cumulative number of confirmed cases and deaths.

The COVID-19 epidemic impact on the regional and global economy has influenced decisions on how and when businesses, public centers, tourism, schools and universities can safely reopen [23]. For decision-makers it is important to count with a quantitative evaluation of the measures that were taken and those that could have been taken during the SARS-CoV-2 pandemic in order to design and apply better and more effective mitigation strategies in the likely event of a new infectious disease emergency. This knowledge is even more pressing in countries that lack the full infrastructure to acquire a more precise or, perhaps we should say, a less uncertain idea of the behavior of the pandemic. This paper attempts to provide such evaluation.

The manuscript is organized as follows. In Section 2, we present the formulation of the mathematical model and the methodology for parameter estimation. In Section 3, we show the results of the model fitting for Mexico and Peru. Section 4 makes a retrospective analysis considering hypothetical interventions that occurred on predefined key dates. Finally, discussion and conclusions can be found in Section 5.

## 2. Methods

### 2.1. Mathematical model

A compartmental model is used to describe the evolution of the COVID-19 pandemic. The model considers three classes of infected individuals: Asymptomatic (I), Symptomatic (Y) and Reported (T). Once reported, infected individuals are effectively isolated and no longer participants in the transmission process. The model allows Susceptible (S) individuals to be Vaccinated (V) with a vaccination rate *ψ*. It is assumed that the vaccine is not perfect which implies that vaccinated people can be infected. Likewise, it is assumed that vaccinated individuals become susceptible after a certain period. Vital dynamics are also included since this work models the first year of the pandemic. Figure 1 shows the corresponding model diagram.

**Figure 1.**
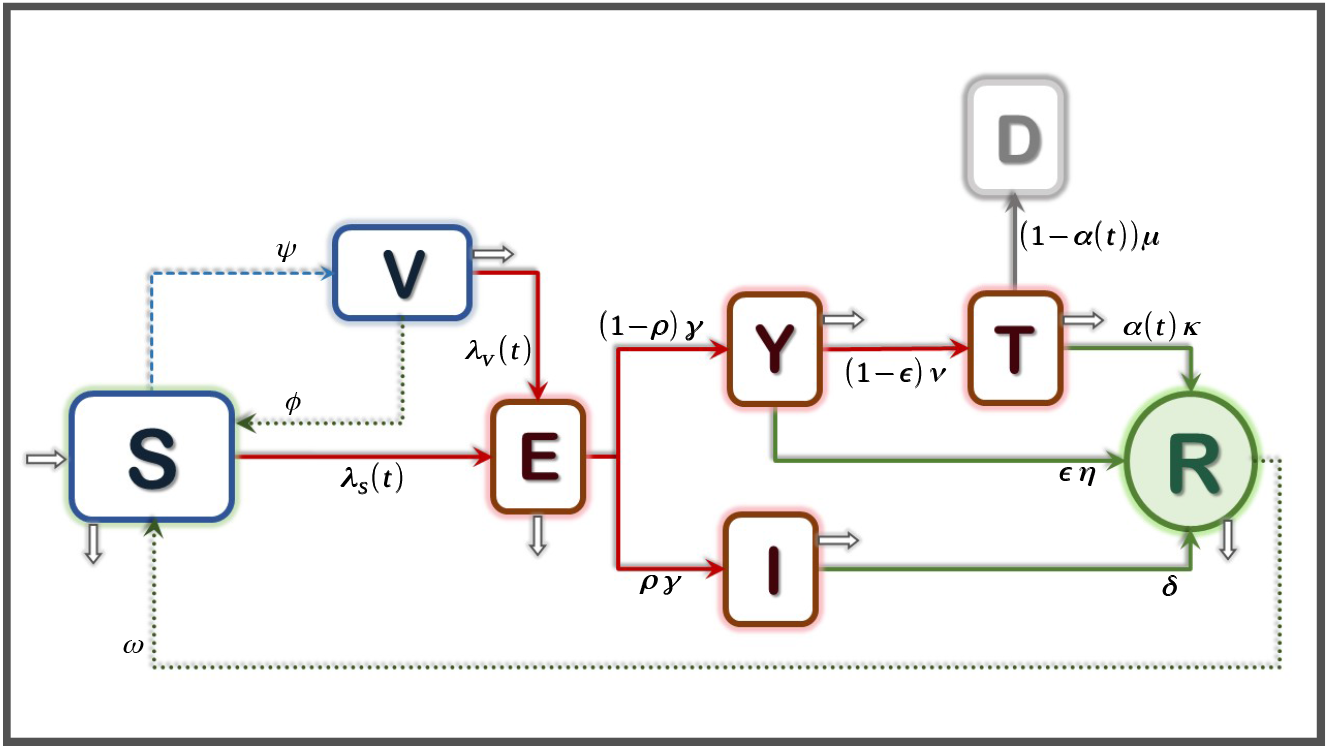
Mathematical model diagram. There are three types of infectious individuals: asymptomatic, symptomatic and reported. Reported cases do not play a role in transmission. Here, *λ*_*S*_(*t*) and *λ*_*V*_ (*t*) represent the infection force related to susceptible and vaccinated people, respectively.

The mathematical model is given by the system of ordinary differential equations shown in the system (2.1).

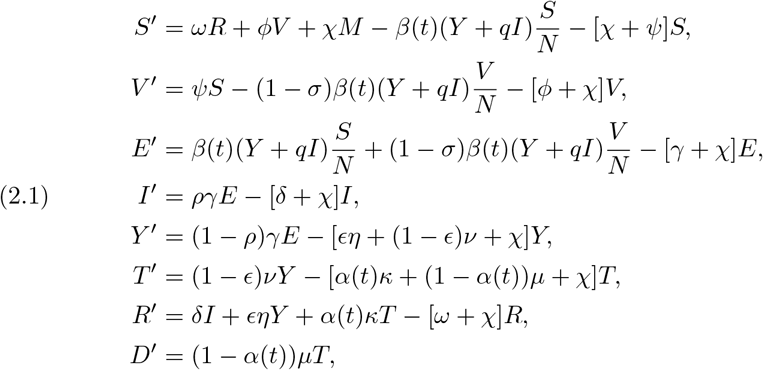

Note that, in (2.1), *N* = *S* + *V* + *E* + *I* + *Y* + *R* and *M* = *N* + *T*. Table 1 shows a description of all the model’s parameters and their values.

**Table 1.** Description and values of the model’s parameters given in System 2.1. See [1, 28] for sources.

| Parameter | Description | Value | Units |
| --- | --- | --- | --- |
| $\psi$ | Vaccination rate | varying | days<sup>-1</sup> |
| $\nu$ | Screening rate | varying | days<sup>-1</sup> |
| $\mu$ | Disease mortality rate | varying | days<sup>-1</sup> |
| $\phi$ | Vaccine immunity waning rate | 0.006 | days<sup>-1</sup> |
| $\omega$ | Natural immunity waning rate | 0.006 | days<sup>-1</sup> |
| $\gamma$ | Incubation rate | 0.196 | days<sup>-1</sup> |
| $\delta$ | Asymptomatic recovery rate | 0.143 | days<sup>-1</sup> |
| $\eta$ | Symptomatic recovery rate | 0.071 | days<sup>-1</sup> |
| $\kappa$ | Reported recovery rate | 0.1 | days<sup>-1</sup> |
| $\chi$ | Natural mortality rate | $3.629 \times 10^{-5}$ | days<sup>-1</sup> |
| $\rho$ | Proportion of asymptomatic | 0.35 | % |
| $\epsilon$ | Proportion of symptomatic recovered | 0.94 | % |
| $q$ | Asymptomatic infectiousness reduction | 0.45 | % |
| $\sigma$ | Vaccine efficacy | varying | % |
| $\alpha(t)$ | Proportion of reported recovered people | estimated | % |
| $\beta(t)$ | Effective transmission contact rate | estimated | days<sup>-1</sup> |

The model depicted in Figure 1 is standard, but its main feature is how the contact rate *β* and the proportion of reported recovered people *α* are handled. Both parameters are time-dependent and defined through the interpolation of *k* change points. Interpolation is done using Hermite polynomials instead of splines to guarantee that both rates remain positive at any point in time. It is possible to estimate these two functions by using two time series: the number of reported cases and deaths. The *k* points in time when changes in *β* and *α* occur are predefined dates associated with the beginning of the mitigation measures and civic or religious holidays.

#### 2.1.1. Vaccination parameters

Many countries worldwide are using more than one vaccine. However, our model does not incorporate a detailed vaccination dynamic with different vaccines, doses, and efficacy since there is limited information regarding this process in Mexico and Peru.

The lack of information on the vaccination process affects the knowledge of important parameter values. Thus, as a first approach, it is assumed that susceptible individuals are vaccinated at a rate *ψ* to estimate the vaccination rate. Since the probability of having been vaccinated at time *t* is 1 − exp(−*ψt*) then, if a proportion *p* of the susceptible population is already vaccinated at time *T*_*V*_, then the vaccination rate that achieves this coverage is given by

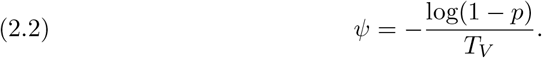

Parameters *p, T*_*V*_ and therefore, *ψ*, will vary between regions depending on the start of vaccination and vaccine stock. More details about the above will be provided in the Section 3.

### 2.2. Parameter estimation

#### 2.2.1. Data

We use the available COVID-19 data of Mexico and Peru [9–11, 15]. Data consists of daily records of reported cases for Mexico, confirmed cases for Peru, and deaths for both countries. We considered data from the start of the pandemic (late February to early March, depending on the specific location) until June 16, 2021.

#### 2.2.2. Statistical inference

A Bayesian approach is used to estimate key parameters of system 2.1: the time dependent contact rate *β*(*t*) and the proportion of recovered reported individuals *α*(*t*).

To simplify the estimation process of functions *β*(*t*) and *α*(*t*), it is assumed that each of them is determined by their values at preset times *τ*_1_, *τ*_2_, …, *τ*_*k*_, which are associated to the key dates of the studied regions. Let *a*_*i*_ be the proportion of reported recovered individuals at time *τ*_*i*_, and *b*_*i*_ the contact rate at time *τ*_*i*_, for *i* = 1, …, *k*. Values of *β*(*t*) and *α*(*t*) for any other point in time are obtained by interpolation using Hermite polynomials instead of splines to guarantee that both rates remain positive.

Let ***θ*** = (*a*_1_, *a*_2_, …, *a*_*k*_, *b*_1_, *b*_2_, …, *b*_*k*_) the vector of parameters that will be estimated. All the other parameters needed to solve system 2.1 are fixed and their values can be found in Table 1. It is assumed that parameters *b*_*i*_ are positive and parameters *a*_1_ must take values in (0,1).

Let *W*_*j*_ and *X*_*j*_ be the random variables that count the number of daily COVID-19 reported infected individuals and deaths at time *t*_*j*_, respectively, for *j* = 1, 2, …*n*. Here, *t*_*j*_ represents the number of days since the start of the pandemic in each region. It is assumed that the probability distribution of *W*_*j*_, conditional on the vector of parameters ***θ***, is a Poisson distribution such that *E*[*W*_*j*_] = *C*(*t*_*j*_|***θ***) −*C*(*t*_*j*−1_ |***θ***), with *C*(*t*|***θ***) being the cumulative number of reported cases according to System 2.1. Similarly, the probability distribution of *X*_*j*_, conditional on the vector of parameters ***θ***, is a Poisson distribution such that *E*[*X*_*j*_] = *D*(*t*_*j*_|***θ***) −*D*(*t*_*j*−1_|***θ***), with *D*(*t* ***θ***) being the cumulative number of deaths according to the compartmental model. Assuming that variables *W*_1_, *W*_2_, …, *W*_*n*_ and *X*_1_, *X*_2_, …, *X*_*n*_ are conditionally independent, then the likelihood function is given by

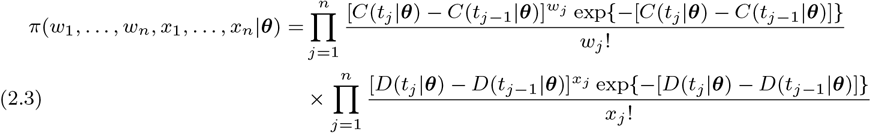

The joint prior distribution for vector ***θ*** is a product of independent uniform distributions and independent log-normal distributions. For parameters *a*_1_, …, *a*_*k*_, the prior is Uniform(0,1), and for *b*_1_, …, *b*_*k*_ the prior is log-normal(0.3, 2). Then

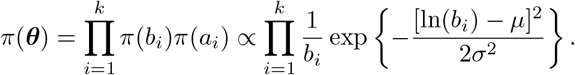

The posterior distribution of the parameters of interest is

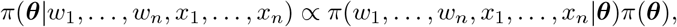

and it does not have an analytical form since the likelihood function depends on the numerical solution of the ODE System 2.1. We analyze the posterior distribution using an MCMC algorithm called *t-walk* [12]. For each region, 3 chains of 8 million iterations are run, from which 200,000 are discarded as burn in. At the end, only 1000 iterations are retained to create the estimations presented in this work.

Of course, confirmed COVID-19 cases can be grouped in different forms in order to represent the epidemic curve. In the case of Mexico, the number of cases can be grouped by date of symptoms onset, by the date when individuals seek medical attention (or tests), and by date of test results. In the case of Mexico, reported cases *X*_1_, …, *X*_*n*_ are those who seek medical attention at time *t*. This is why parameter *ν* is referred as the screening rate, the time from symptoms onset to testing. In the case of Peru, reported cases *X*_1_, …, *X*_*n*_ are those that got positive test results at time *t*. In that case, *ν*^−1^ represents the time from symptoms onset to test results. Notice that there is no need to modify the compartmental model to handle the difference between these two types of data, it is enough to change the parameter *ν*, although the interpretation of the model is slightly different.

## 3. Results

The estimation of the time-varying contact rate *β*(*t*) is central to our work. We first fit the observed cases for both Mexico and Peru. The estimated function *β*(*t*) constitutes the baseline on which comparisons are made when evaluating hypothetical interventions. Figure 2 illustrates the baseline time-dependent contact rate for Mexico City and Lima, estimated from the open repositories of incidence data available in each country. The perturbations that will be described in the following subsections are local, i.e., only change the contact rate on an specific time interval. For each perturbation, outside this interval the rest of the contact rate remains equal to the baseline *β*(*t*) plotted in Figure 2.

**Figure 2.**
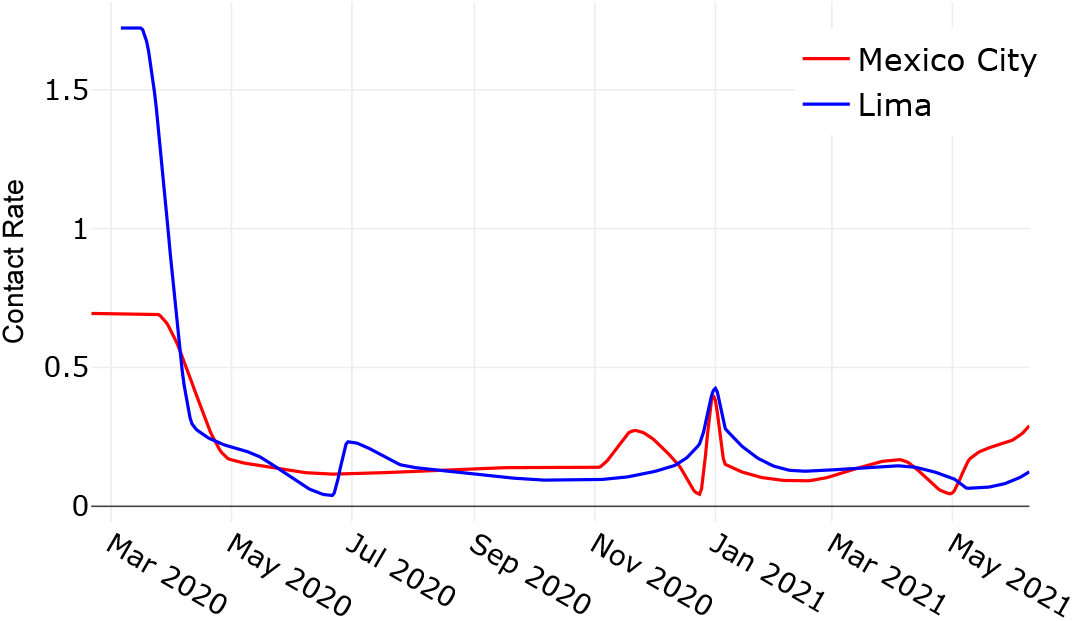
Baseline time-dependent contact rate for Mexico City (red) and Lima (blue).

### 3.1. COVID-19 dynamics in Mexico

As mention above, we use the available COVID-19 data of each Mexican state [15]. The dates when parameters *β* and *α* are assumed to change are shown in Table 2. These dates are the same for all states except for the start of the pandemic.

**Table 2.** Key-dates used to estimate the contact rate and the proportion of reported recovered people for Mexican States.

| Description | Dates | Description | Dates |
| --- | --- | --- | --- |
| First case | Varying | Day of the virgin | 2020-12-12 |
| NPIs start | 2020-03-23 | Christmas | 2020-12-24 |
| Childrens day | 2020-04-30 | New Year | 2020-12-31 |
| Mothers day | 2020-05-10 | Wise men day | 2021-01-06 |
| Fathers day | 2020-06-21 | Valentines day | 2021-02-14 |
| Independence day | 2020-09-16 | Easter | 2021-04-04 |
| Day of the dead | 2020-11-02 | Childrens day | 2021-04-30 |
| Buen Fin ends | 2020-11-21 | Mothers day | 2021-05-10 |

To calculate the vaccination rate in Eq 2.2, let, for each state, *T*_*V*_ be the number of days from the start of vaccination up to June 16, 2021. The proportion of vaccinated people *p* during that period is approximated from official communications of the Mexican government [14]. On the other hand, we consider the total number of vaccines applied until June 16, 2021 [14], and the reported efficacy of the vaccines used in Mexico [8] to calculate the weighted vaccine efficacy (*σ*).

Figure 3 shows the COVID-19 model dynamics of reported cases in comparison with the observed data from the beginning of the pandemic to June 9, 2021. To exemplify the results, we chose states located at the north, center, and south of the country: Sonora, Mexico City, Queretaro, and Quintana Roo. Note that each epidemic curve has a different behavior. Nevertheless, the proposed scheme based on key dates provides a good fit for the data.

**Figure 3.**
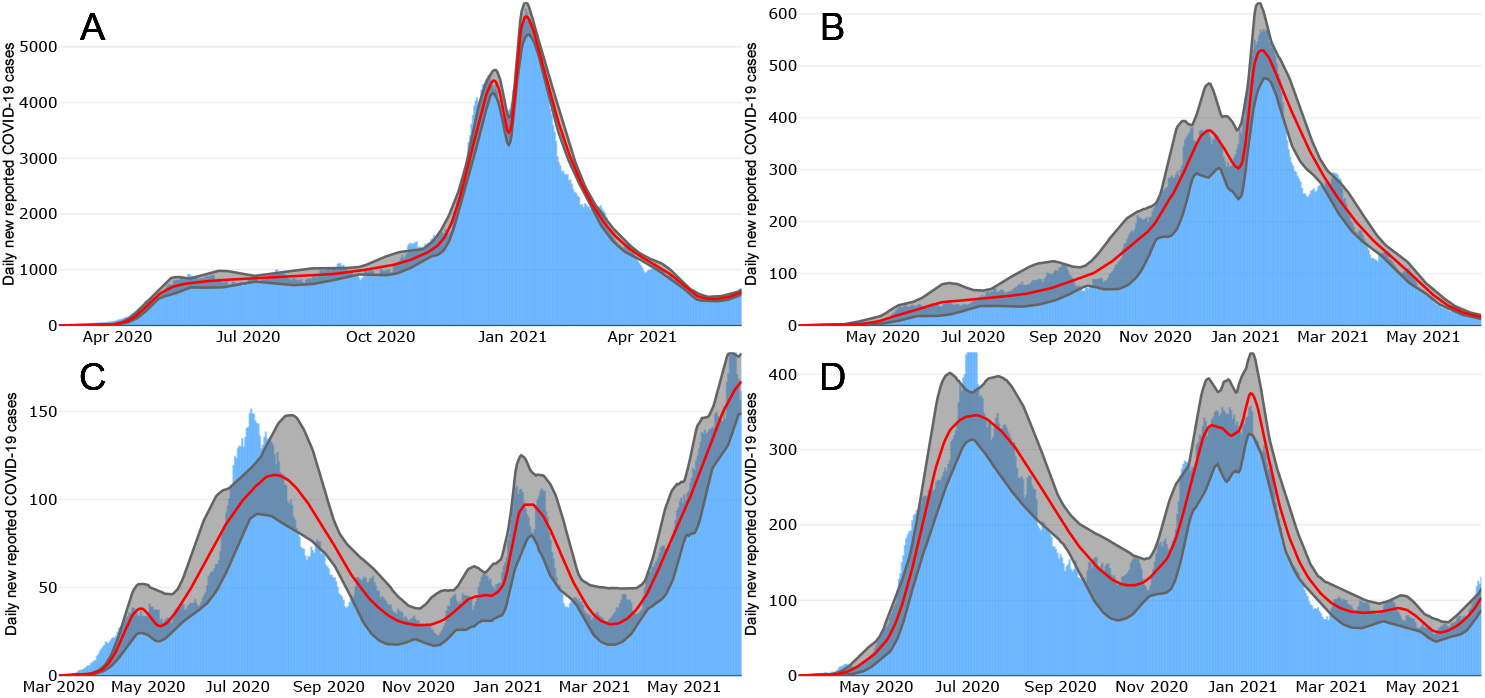
New reported COVID-19 cases in four Mexican states. (A) Mexico City, (B) Queretaro, (C) Sonora, and (D) Quintana Roo. Blue bars show reported cases data. Red lines show the median posterior estimates. The gray shadow represents 95% pointwise probability regions.

Figure 4 shows the daily mortality given by the model in comparison with the observed deaths until June 9, 2021 for the same Mexican sates. The fit is also good even when the observed data shows state specific patterns. Furthermore, observe that the dynamics of reported cases and deaths is typical to each state.

**Figure 4.**
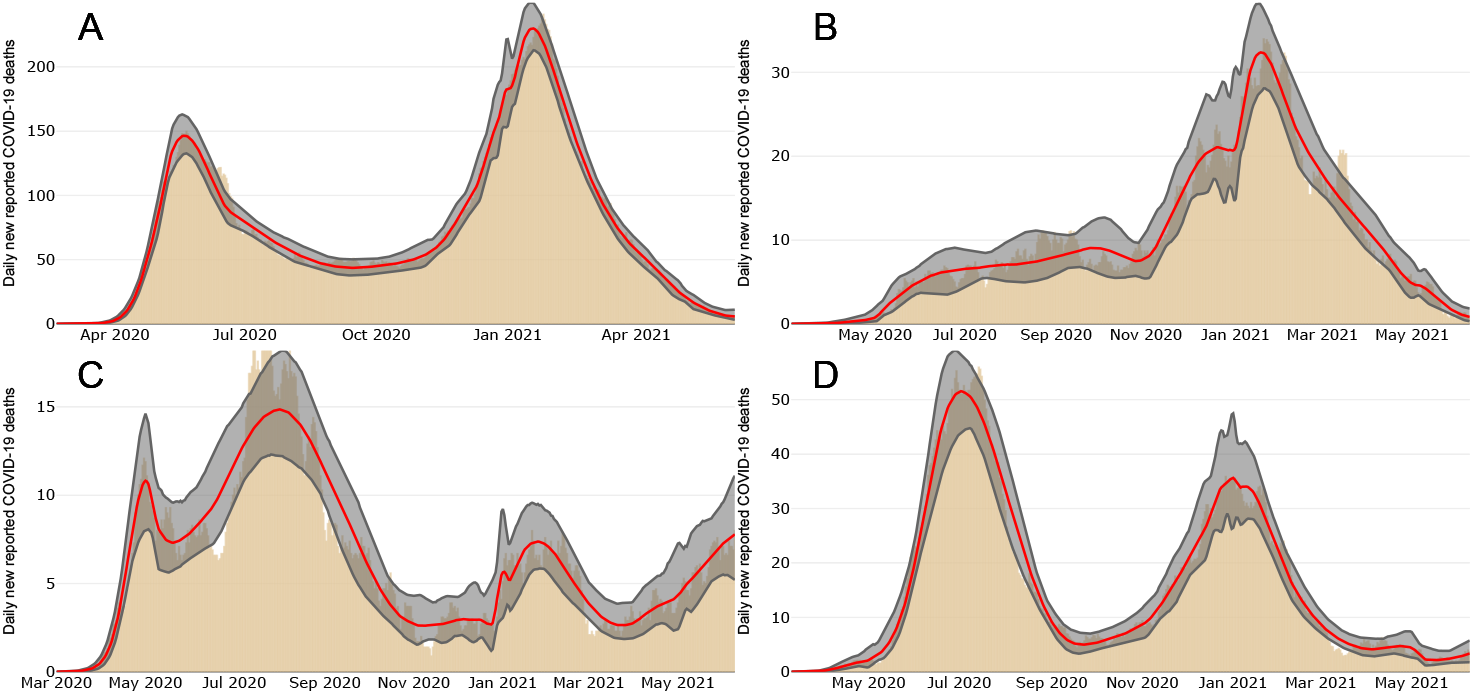
New reported COVID-19 deaths in four Mexican States. (A) Mexico City, (B) Queretaro, (C) Sonora, and Quintana Roo. Black bars show reported deaths. Red lines show median posterior estimates. The gray shadow represents 95% pointwise probability regions.

### 3.2. COVID-19 dynamics in Peru

Using COVID-19 data from Peru from the beginning of the epidemic until June 16, 2021 [9, 10], the corresponding key dates used to estimate parameters *β* and *α* for Peru are shown in Table 3. Data used to calculate values for *ψ* and *σ* can be found in [11].

**Table 3.** Key-dates used to estimate the contact rate and the proportion of reported recovered people by department (political unit) of Peru.

| Description | Dates | Description | Dates |
| --- | --- | --- | --- |
| First case | Varying |  |  |
| NPIs start | 2020-03-16 | Saints' day | 2020-11-01 |
| Easter | 2020-04-12 | Immaculate Conception day | 2020-12-08 |
| Labor day | 2020-05-01 | Christmas | 2020-12-24 |
| Mothers day | 2020-05-10 | New Year | 2021-01-01 |
| Fathers day | 2020-06-21 | Wise men day | 2021-01-06 |
| St Peter and St Paul day | 2020-06-29 | Valentines day | 2021-02-14 |
| Independence day | 2020-07-28 | Easter | 2021-04-04 |
| Santa Rosa de Lima day | 2020-08-30 | Labor day | 2021-05-01 |
| Battle of Angamos | 2020-10-08 | Mothers day | 2021-05-09 |

Figures 5 and 6 show observed and fitted COVID-19 confirmed cases and mortality, respectively, in four departments of Peru: Arequipa, Cusco, Lima and Piura.

**Figure 5.**
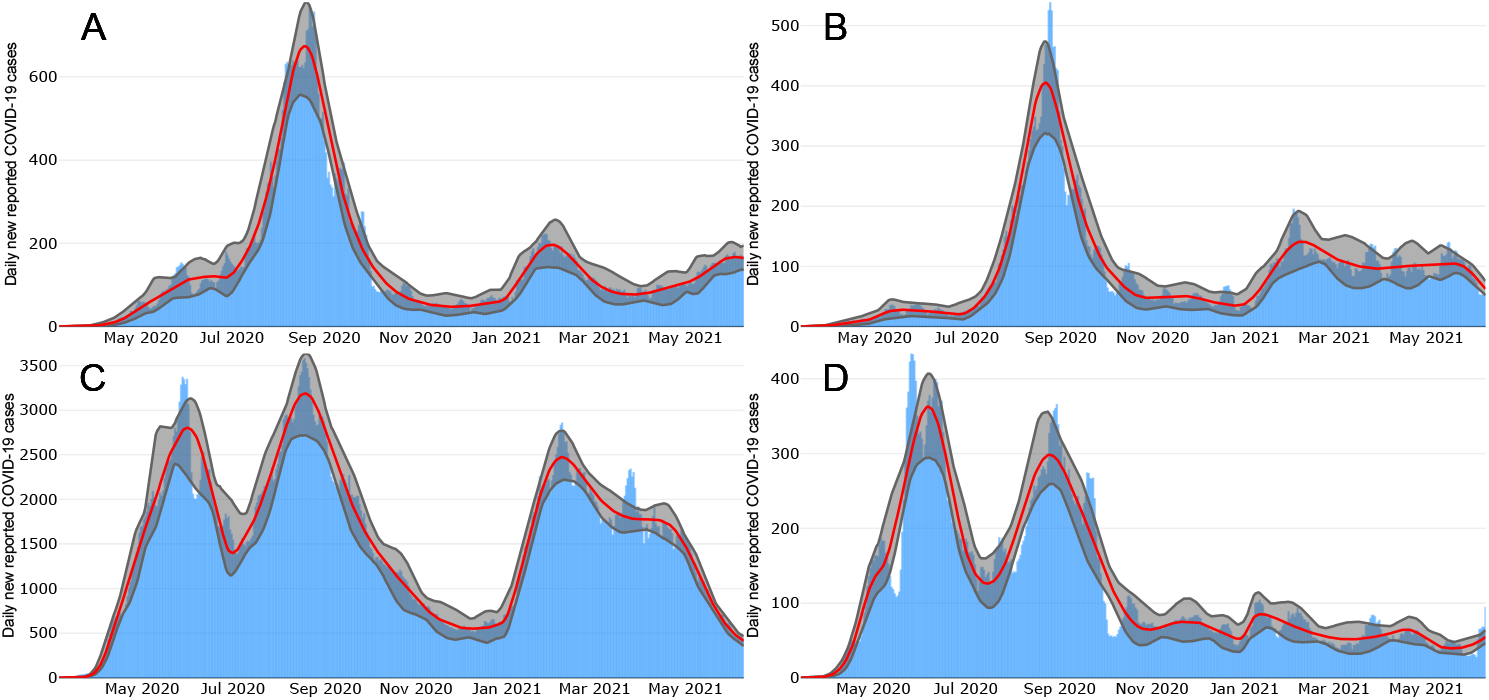
New confirmed COVID-19 cases in four departments of Peru. (A) Arequipa, (B) Cusco, (C) Lima, and (D) Piura. Blue bars shows confirmed cases. Red lines show the median posterior estimates. The gray shadow represents 95% point-wise probability regions.

**Figure 6.**
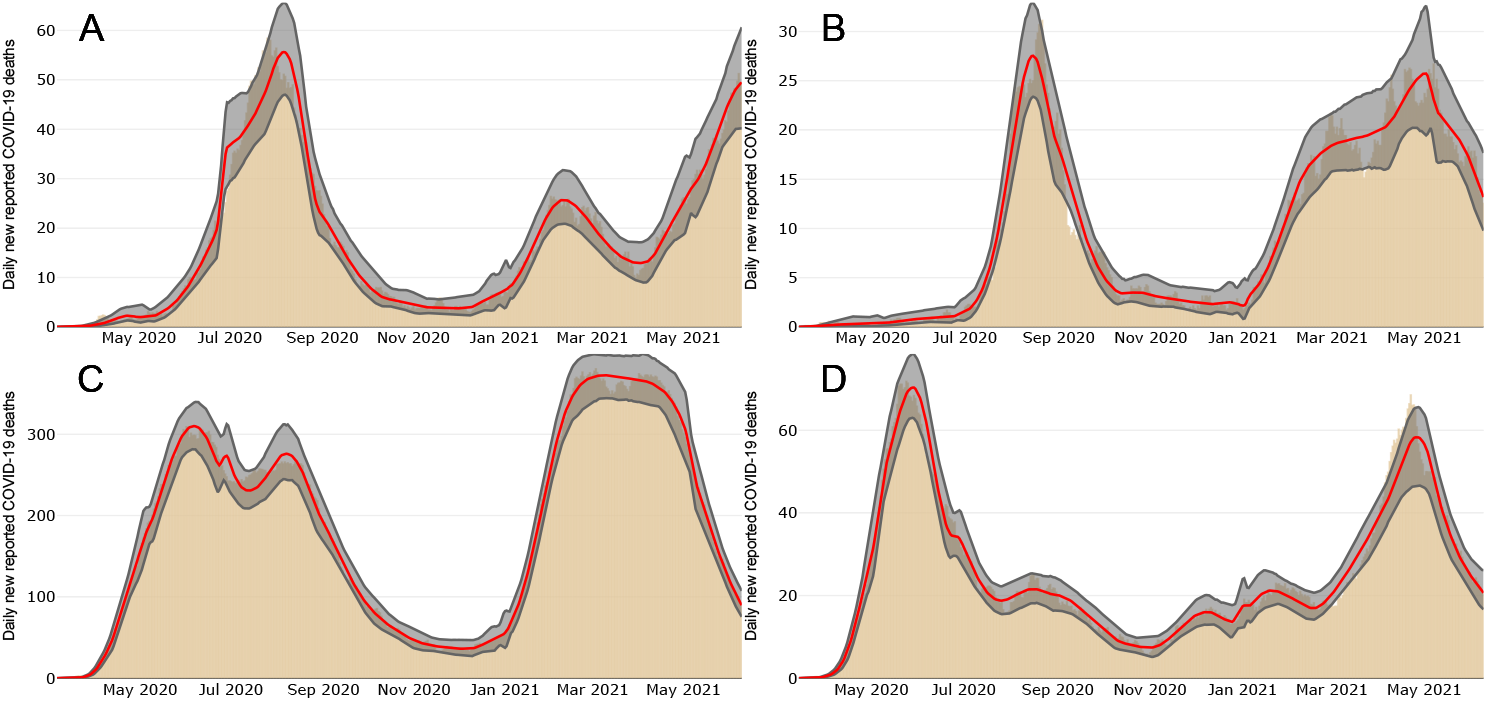
New reported COVID-19 deaths in some departments of Peru. (A) Arequipa, (B) Cusco, (C) Lima, and Piura. (D) Beige bars shows reported cases data. Red lines show the median posterior estimates. The gray shadow represents 95% pointwise probability regions.

We can see that, despite obvious differences in the shape of the epidemic curve between both countries, the fit continues to be good for confirmed cases and deaths.

## 4. Looking back: hypothetical trends under different interventions

As we know, at the beginning of the COVID-19 pandemic, countries around the world implemented different mitigation measures to control COVID-19 disease transmission. However, these measures were largely insufficient to control disease contagion and mortality in many places. A high mortality rate and the occurrence of more than one outbreak in the first year of the pandemic are evidence of this fact. As we have argued before, key calendar dates are civic, religious or commercial days where transmission was observed to increase because of the relaxation or lack of enforcement of NPIs indicatives. To explore the impact of stricter NPIs executed or enforced during key calendar dates, we use our mathematical model. We evaluate the hypothetical epidemic trends obtained by the reduction of the contact rate at different strengths and different key dates. Contact rate perturbations are continuous and are defined on an interval of successive key dates in a continuous way. The percentage reduction that we report refers to the maximum reduction achieved in this interval. Changes in the contact rate are applied in three periods during the first year of the pandemic: (i) near the beginning of the pandemic, (ii) at the beginning of the second outbreak, and (iii) at the end of the year.

In the following we present our results arising from making local changes in the contact rate at the times listed above. Recall that, for our scenarios, except for the local perturbation in each case, the rest of the contact rate is the same as in the baseline case. This assumes that the general social distancing behavior and compliance with NPIs of the population observed during 2020, expressed in the magnitude and trend of the contact rate, does not change except on the perturbed key dates (scenarios (i), (ii), and (iii) above).

### 4.1. Mexico City

Figures 7 and 8 show Mexico City scenarios regarding newly reported COVID-19 cases and deaths, respectively. Gray, blue, and green lines show COVID-19 dynamics when reducing the value of transmission contact rate to 25%, 50%, and 75%, respectively. Red line represents our baseline COVID-19 dynamics. We can observe that both figures evidence similar trends in their behaviors.

**Figure 7.**
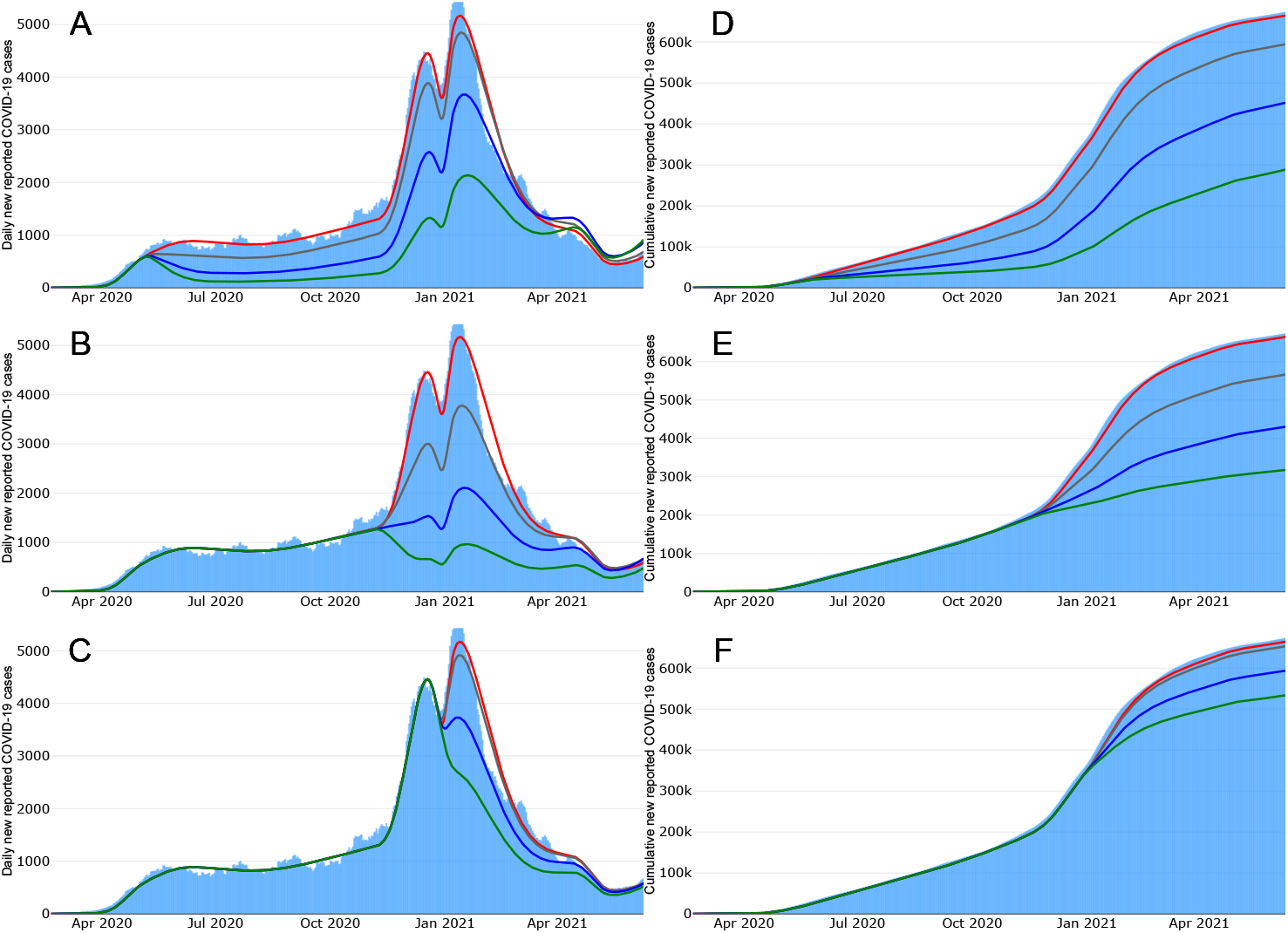
Mexico City Scenarios - New reported COVID-19 cases. Upper and lower rows represent the daily and cumulative reported COVID-19 cases, respectively. Panels A and D illustrate COVID-19 dynamics when perturbing the transmission contact rate on Mother’s day. Panels B and E show COVID-19 dynamics when perturbing the transmission contact rate by the end of the *Buen Fin*. Panels C and F illustrate COVID-19 dynamics when perturbing the transmission contact rate on New Year’s day.

**Figure 8.**
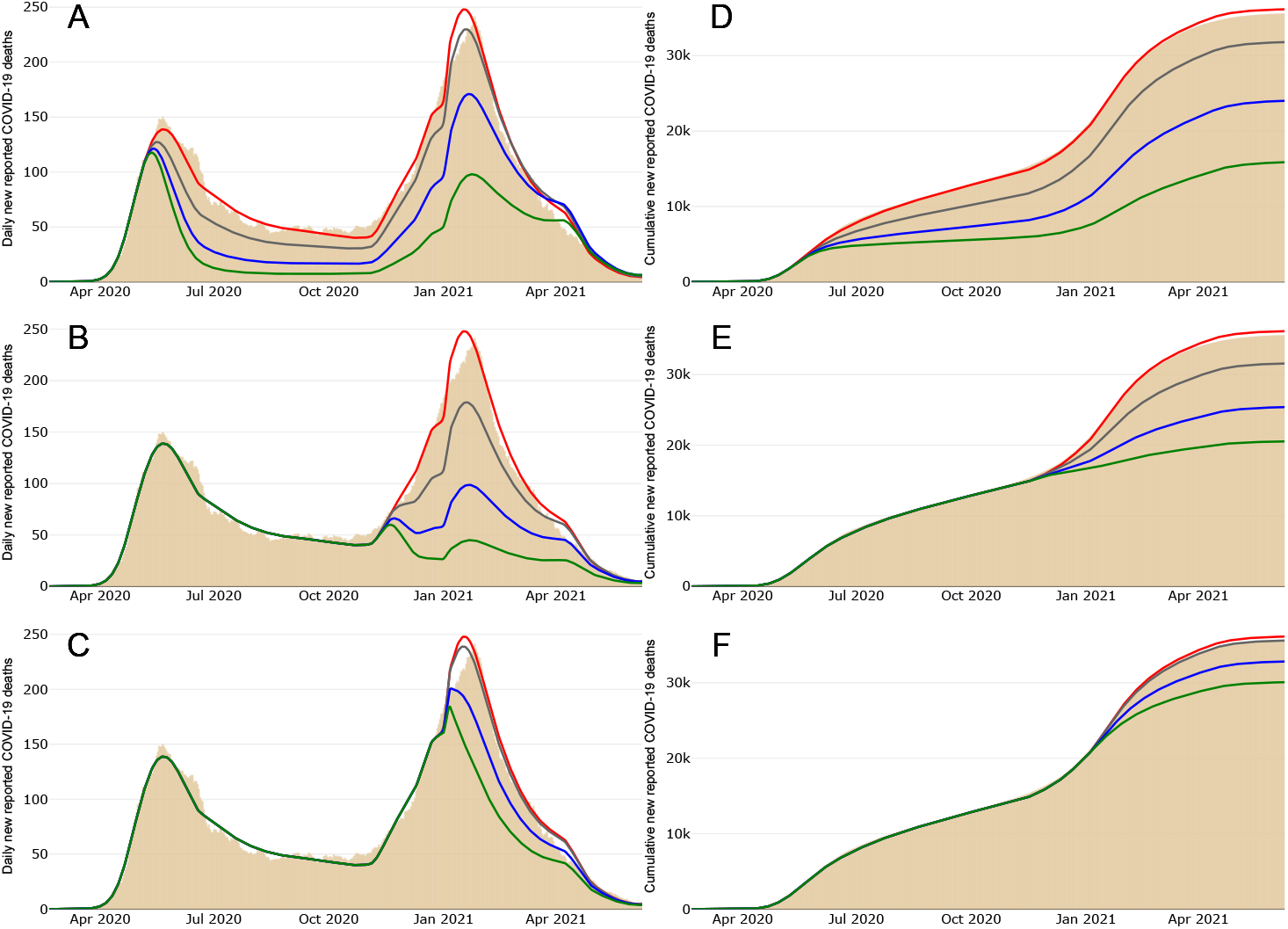
Mexico City Scenarios - New reported COVID-19 deaths. Upper and lower rows represent the daily and cumulative reported COVID-19 deaths, respectively. Panels A and D illustrate COVID-19 dynamics when perturbing the transmission contact rate on Mother’s day. Panels B and E show COVID-19 dynamics when perturbing the transmission contact rate by the end of the *Buen Fin*. Panels C and F illustrate COVID-19 dynamics when perturbing the transmission contact rate on New Year’s day.

Figure 7A shows that all reductions in case (i) lead to a period of slow growth longer than observed in the baseline. We have argued before [28] that this is due to the proximity of the intervention day to the peak of the epidemic that occurred in the last week of May. With respect to outbreaks, all contact rate reductions generate outbreaks with lower intensity than the baseline. Moreover, taking June 9, 2021 as the final date of our analysis, the most extreme intervention (75% reduction) produces a reduction close to 56% of the baseline cumulative incidence (see Figure 7D).

Figure 7B illustrates that reductions in case (ii) occur at a period of epidemic growth far from the observed peak. For this case the reductions on incidence are significant; in particular, the most extreme intervention (75% reduction), the incidence changes direction and takes a downward trend. However, if we consider June 9, 2021, as the final date of our analysis, the most extreme intervention in case (ii) is less effective than in case (i). Cumulative incidence presents a reduction close to 52% regarding the baseline dynamics (see Figure 7E). This last may be a consequence of the length of the period perturbation in case (i) being greater than in case (ii).

Finally, Figure 7F shows that acting on a period coincident with the peak of the curve (case (iii)) achieves comparatively poor reductions in the cumulative incidence even in the case of the most extreme intervention (where the reduction is close to 19% of the baseline cumulative incidence). However, Figure 7C shows that both medium and extreme interventions notably reduce second-peak levels of incidence which is very useful to avoid the health centers’ saturation.

### 4.2. Lima

Figures 9 and 10 show Lima scenarios regarding new confirmed COVID-19 cases and new reported COVID-19 deaths, respectively. Gray, blue, and green lines show COVID-19 dynamics when reducing the value of transmission contact rate to 25%, 50%, and 75%, respectively. Red line represents our baseline COVID-19 dynamics. We can observe that both figures evidence similar trends in their behaviors.

**Figure 9.**
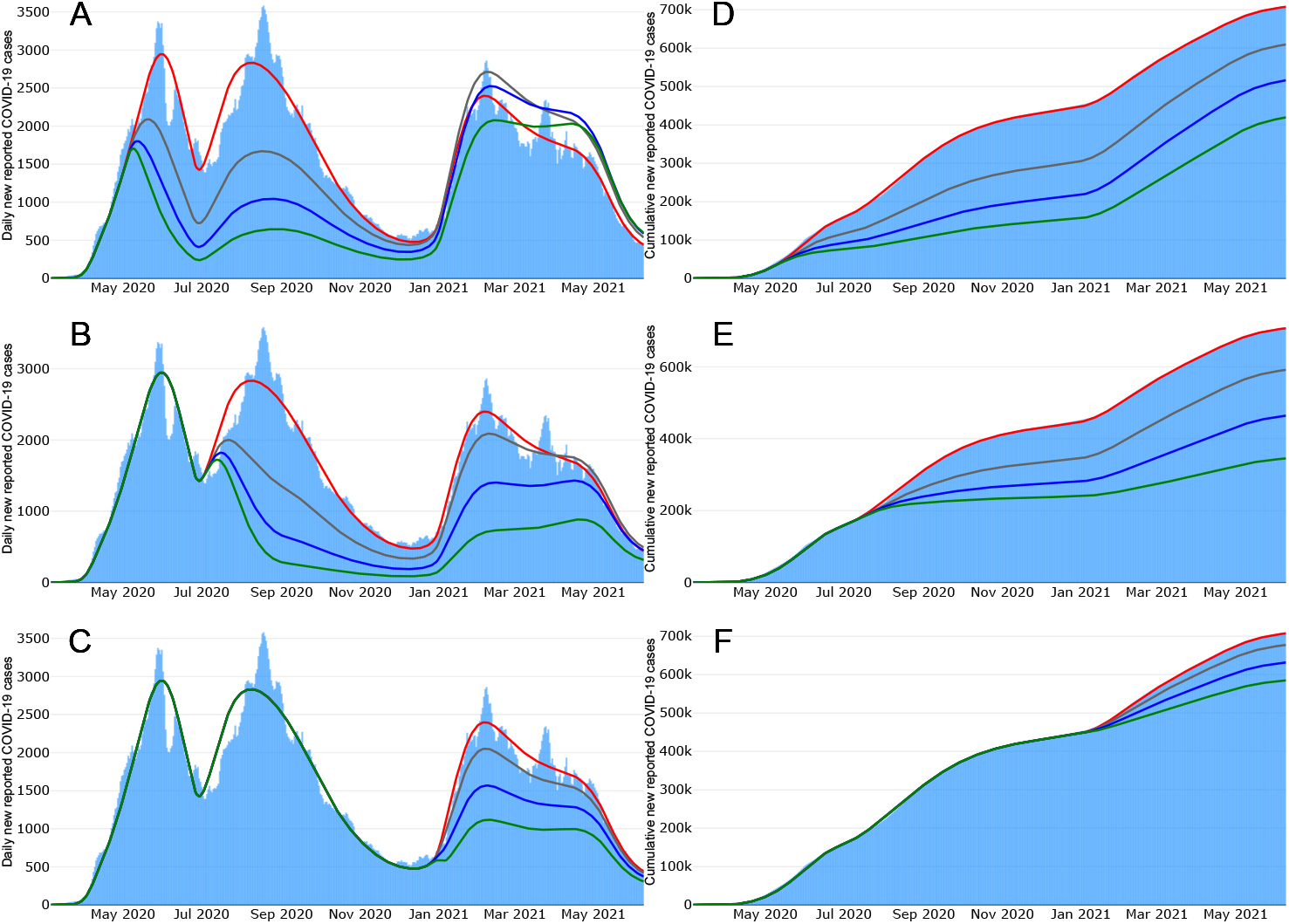
Lima Scenarios - New confirmed COVID-19 cases. Upper and lower rows represent the daily and cumulative confirmed COVID-19 cases, respectively. Panels A and D illustrate COVID-19 dynamics when perturbing the transmission contact rate on Mother’s day. Panels B and E show COVID-19 dynamics when perturbing the transmission contact rate on Independence day. Panels C and F illustrate COVID-19 dynamics when perturbing the transmission contact rate on New Year’s day.

**Figure 10.**
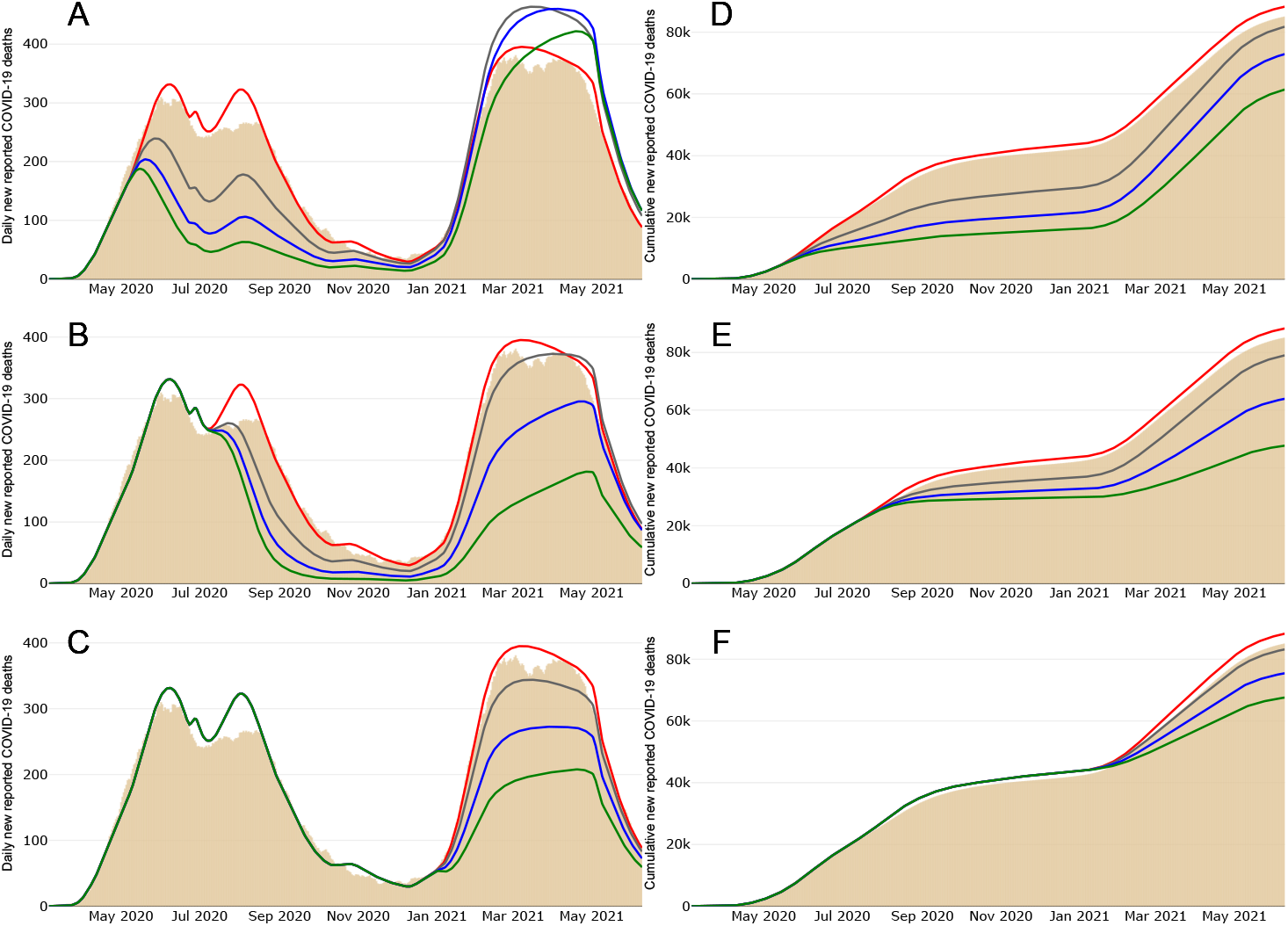
Lima Scenarios - New reported COVID-19 deaths. Upper and lower rows represent the daily and cumulative reported COVID-19 deaths, respectively. Panels A and D illustrate COVID-19 dynamics when perturbing the transmission contact rate on Mother’s day. Panels B and E show COVID-19 dynamics when perturbing the transmission contact rate on Independence day. Panels C and F illustrate COVID-19 dynamics when perturbing the transmission contact rate on New Year’s day.

Figure 9 shows the COVID-19 epidemic curve for Lima. It certainly differs from that observed in Mexico City. Even though a curfew was imposed, there was more than one outbreak at the beginning of the pandemic. This indicates that the curfew did not achieve the intended aim which was to reduce transmission hinting, perhaps, of a defficient enforcement and compliance. The case of China shows that efficient curfews can be very effective in suppressing transmission.

In contrast to Mexico City, Figure 9A shows that case (i) results in a different trend for Lima. The intervention occurs on a date far from the observed peak and, therefore, similarly to case (ii) in Mexico, the impact of these interventions reduces the nearby outbreaks notably. Moreover, as in case (i) for Mexico, acting early does reduce cumulative incidence overall (see Figure 9D). However, we also observe that all early reductions resulted in larger peaks on the third outbreak. This is likely a consequence of a larger susceptible pool associated with early interventions.

Figure 9B illustrates that, in Lima, case (ii) happens when the epidemic curve is in a downward trend and reaches a local minimum. The consequence of the intervention at this time is an effective reduction of the size of the later peak.

Moreover, on June 9, 2021, the final date of our analysis, the most extreme intervention (75% reduction) generates a reduction close to 51% of the baseline cumulative incidence (see Figure 9E).

Figures 9C and F show that, as in the case of Mexico, case (iii) evidences a poor reduction in prevalence even for the most strict of the reductions.

Moreover, on June 9, 2021, the final date of our analysis, the most extreme intervention generates a reduction close to 17% of the baseline cumulative incidence (see Figure 9F). However, we also observe that all reductions generate a decrease in the incidence, that produce a plateau behavior for the most extreme intervention.

## 5. Discussion and Conclusions

Emerging infectious diseases are an important concern for public health. COVID-19 disease is the more recent example that has caused more than 6.6 million deaths and 650 million confirmed cases around the world as of middle December, 2022 [22]. This disease has once again shown the role of mobility in the spread of acute respiratory diseases [28]. This is consistent with research where superspreading events are found to be [21].

The epidemic dynamics observed in all of Mexico have been driven by events associated with heightened mobility and increased social activity [2,5,20,25,26,31, 34, 36] during holidays and other important calendar key dates. The older population as well as that with comorbidities, such as obesity, diabetes, and hypetension, has been most affected [27]. Early in the pandemic, key calendar days as described here, were associated with large gatherings violating the social-distancing policies in effect at the time. These dates provide good support to putative changes in the contact rates. As an example, in Mexico City the first case occurred by the end of February 2020, and the first set of mitigation measures was applied on March 23, 2020. Later that year, there were superspreading events in Mexico City during Easter celebrations (April 6-12) and early May (April 30 to May 10) that shifted the day of maximum incidence to the end of May, and pushed the epidemic into a quasi-stationary state characterized by values of *R*_*t*_ ≈1 [1, 28]. This behavior, observed around the world, has been explored in [18, 29, 32]. In particular, [32] claim that this quasi-linear growth and the maintenance of the effective reproduction number around *R*_*t*_ ≈ 1 for sustained periods of time, involves critical changes in the structure of the underlying contact network of individuals. In the case of Mexico City, we have argued [28] that these changes relate to superspreading events on key calendar dates (Easter holidays and children’s, Holy Cross’ and Mother’s days).

This study extends the ideas presented in [28]. We develop a methodology for the description of the epidemic curve based on key calendar days that allows us to recreate the COVID-19 dynamics in different cities. Our main hypothesis assumes that key calendar days are associated to events where the contact rates change and, therefore, are central points to estimate parameters and explain the observed epidemic dynamics. We employed the same methodology for Mexico and Peru obtaining good results both for incidence and death dynamics (see Figures 3 to 6) showing that our approach can recreate different types of COVID-19 dynamics and likely can be extended to other acute respiratory infections.

Using our model and the associated methodology, we explore the impact of hypothetical interventions that reduce the contact rate in a given period. As expected, all reductions decrease the cumulative incidence. The magnitude of this reduction will depend on the magnitude of the reduction of the contact rate. For both Mexico and Peru, reductions of the contact rate early in the epidemic are enough to lower the prevalence for the whole year. However, in Peru, an early reduction can also generate the opposite effect later in the year since our simulations show that the outbreak in the first quarter of 2021 is always greater than the baseline. This phenomenon can be explained by the existence of a large susceptible pool generated by the early intervention on the contact rate. On the other hand, a reduction of the contact rate at the end of the first year produces a smaller reduction in the cumulative incidence, but will produce a significant reduction in the magnitude of the following, later, outbreak. This effect is positive because it would help to reduce hospitalizations and the saturation of the health centers that was almost reached both in Mexico and Peru in early 2021 [3, 33].

This study has shown that the key dates can be useful for the implementation of non-pharmaceutical interventions (NPIs). Key dates are known, which would help to design a strategy ahead of time. Likewise, we have shown that the enforcement of NPIs will always generate a reduction in the cumulative incidence, which, even if minimal, can greatly impact the magnitude of the epidemic outbreaks. Finally, there are still relevant questions to answer mainly centered on planning strategies in advance. We have shown that prior knowledge of key dates (civil, religious, political) can be useful to provide an accurate description of epidemic trends when testing and contact tracing are not widely available.

## Data Availability

All data produced in the present study are available upon reasonable request to the authors

## Author Contributions

### Conceptualization

Manuel A. Acuña-Zegarra, Mario Santana-Cibrian, Jorge X. Velasco-Hernández.

### Discussion and Draft Preparation

Manuel A. Acuña-Zegarra, Mario SantanaCibrian, Carlos E. Rodriguez Hernandez-Vela, Ramsés H. Mena, Jorge X. Velasco-Hernández.

